# Regional versus Extended Lymph Node Dissection for Intrahepatic Cholangiocarcinoma: a randomized controlled trial protocol

**DOI:** 10.1101/2022.10.07.22280815

**Authors:** Li Qi, Feng Yangyang, Jin Yun, Xi Longfu, Yu Yuanquan, Guo Yinghao, Zhang Xiaoxiao, Wu Di, Li Jiangtao

## Abstract

**Introduction:** The mainstay treatment for intrahepatic cholangiocarcinoma is surgical resection, the impact of lymph node dissection and the scope of lymph node dissection for intrahepatic cholangiocarcinoma surgery is controversial. However, research on the comparative of regional lymphadenectomy and extended lymphadenectomy is still limited. This study will explore the effect of regional lymphadenectomy and extended lymphadenectomy in intrahepatic cholangiocarcinoma surgery patients to provide reliable evidence for further standardize the scope of lymph node dissection for intrahepatic cholangiocarcinoma surgery.

**Methods and analysis:** This is a prospective, multicentre, randomised controlled trial in intrahepatic cholangiocarcinoma surgery patients. A total of 174 patients will be enrolled at the first clinic visit in sites from China. Participants will be allocated randomly to the regional lymphadenectomy group and extended lymphadenectomy group (1:1 ratio). Patients in different groups undergo the corresponding surgery. Primary study outcome is disease-free survival (DFS). Secondary study outcomes include incidence of severe Clavien–Dindo complications (≥II), safety of operation, overall survival (OS), three-year survival rate, five-year survival rate and median mortality. Participants will be followed up at baseline, post-operation, every 3 months for the first 2 years and 6 months for the next 3 years to evaluate the impact of different surgery types. The analysis will be done according to protocol cohorts, adjusted by variables associated with intrahepatic cholangiocarcinoma.

**Ethics and dissemination:** This study was approved by the ethical review committee of the second affiliated hospital Zhejiang University school of medicine (2019-261) and will provide reference for standardize the scope of lymph node dissection for intrahepatic cholangiocarcinoma surgery.

**Trial registration:** ClinicalTrials.gov, NCT04078230. Registered on September 6, 2019. The study is expected to last for more than five years, and the first patient was enrolled on August 22, 2020, and a total of 17 patients have been enrolled so far.

## BACKGROUND

Intrahepatic cholangiocarcinoma (ICC) represents 10-15% of liver cancers which is the second most common liver cancer.^[1]^ The incidence of ICC has increased worldwide over the past few decades, some studies suggest a possible link to viral hepatitis, metabolic syndrome and chemical contact.^[2-6]^ Under this trend, there is increasing interest in the standardized treatment of intrahepatic cholangiocarcinoma.

Surgical resection is the only effective treatment for ICC, but the postoperative recurrence rate is high, the median disease-free survival (DFS) is only 20 months, and the 5-year survival rate is about 14-30%. It is a malignant tumor with a poor prognosis. ^[7,8]^ Pathological classification, tumor size and number, lymph node metastasis (LNM) and other factors may be related to the poor prognosis of ICC.^[9-14]^ Several studies have shown that lymph node metastasis is a major risk factor for poor prognosis.^[10,12,15]^

### Strengths and limitations of this study

A major advantage of this trial is that its multicenter, large sample, randomized controlled design will yield reliable results regarding the intervention.

Patients from multiple regions made the study results broadly representative.

The surgeon will inevitably know the procedure that the patient undergoes, and although we exclude the surgeon from the experimental process outside of the operation, it may still have an impact on the results of the experiment.

In previous studies, lymph node metastasis was presumed an intermediary step in dissemination to distant sites.^[16,17]^ But with the deepening of research, new results show that lymph node metastasis plays a more important and complex role in tumor development and metastasis. Primary tumors elicit anti-tumor immune responses by presenting antigens to lymph nodes, which may play an important role in tumor immunotherapy.^[18-21]^ A study in 2022 subversively proposed that lymph node metastases can resist T cell-mediated cytotoxicity induce tumor-specific immune tolerance and lead to distant tumor metastasis.^[22]^

Different opinions on the role of lymph node metastasis in the progression and metastasis of intrahepatic cholangiocarcinoma have triggered controversy on the scope of lymph node dissection during radical resection of cholangiocarcinoma. The American Joint Committee on Cancer (AJCC) 8th edition indicates that radical resection of cholangiocarcinoma should remove at least 6 lymph nodes.^[23]^ American Hepato-Pancreato-Biliary Association (AHPBA) expert consensus statement suggested that the hepatoduodenal ligament, hepatic artery and retropancreatic lymph nodes should be dissected for ICC originating from the right hemiliver; the hepatoduodenal ligament, hepatic artery lymph nodes and nodes around the cardiac portion of the stomach and along the lesser curvature should be dissected for ICC originating from the left hemiliver.^[24]^ NCCN Guideline (V 2.2022) only support a regional lymphadenectomy dissection of the porta hepatis lymph nodes. Other studies proposed to group the lymph nodes according to the tumor location and expand the scope of lymph node dissection. For the right lobe tumor: Group 1 included lymph nodes in the hepatoduodenal ligament; Group 2 included left gastric artery, common hepatic artery, coeliac artery and posterior surface of the pancreas head lymph nodes; Group 3 are distant nodes. Lymph nodes in the right cardial region and along the lesser curvature of the stomach are added to group 1 when the tumor is located in the left lobe.^[25]^

Extended lymphadenectomy can obtain more lymph nodes and improve the accuracy of staging. However, whether this operation can inhibit tumor metastasis and recurrence, prolong the survival time and improve the quality of life of patients is still unclear. Therefore, we will design a multicenter randomized controlled study to assess the effect of extended lymph node dissection on patient prognosis and surgical safety.

The objectives of this trial are: (1) To evaluate whether extended lymph node dissection can improve the prognosis of patients undergoing radical cholangiocarcinoma; (2) To evaluate the safety and reliability of extended lymph node dissection; (3) To evaluate whether extended lymph node dissection can improve the accuracy of staging of intrahepatic cholangiocarcinoma.(4) To standardize the scope of lymph node dissection in radical resection of intrahepatic cholangiocarcinoma.

## METHODS

This is a prospective, multicentre, randomized controlled trial in intrahepatic cholangiocarcinoma surgery patients. The study will be conducted and reported in accordance with Consolidated Standards of Reporting Trials guidelines.^[37]^

### Study settings

The trial will be conducted in China where incidence of intrahepatic cholangiocarcinoma was 7.55 per 100 000 population.^[23,32,38]^ The epidemiological indicators of intrahepatic cholangiocarcinoma in the study site are shown in Table 1. The selected experimental sites have a large geographical span and cover areas with different medical levels (the sites include cities located in the north, south, east, west of China and developed cities and developing cities).

**Table1.** ICC epidemiology in the study sites^[15,23,26-36]^

| Site/<br>country | Incidence | Five-year<br>survival rate | DFS<br>( months ) | OS<br>( months) | Recurrence<br>rate | LNM rate | Main<br>Etiology |
| --- | --- | --- | --- | --- | --- | --- | --- |
| Global | 6/100000 | 14-30% | 20 | 30 | 40-80% | 17-39.1% | HBV |
| China | 7.55/100000 | 9%~20% | 19.3 | 26.1 | 70% | 45-62% | HBV |

ICC , Intrahepatic cholangiocarcinoma ; DFS , Disease-free survival ; OS , Overall survival ; LNM , Lymph node metastasis ;

### Study population

All patients with intrahepatic cholangiocarcinoma who visited designated medical institutions during the study period will be screened to participate in the trial. The detailed information of the enrolled patients was recorded as shown in Table 2.

**Table 2.**
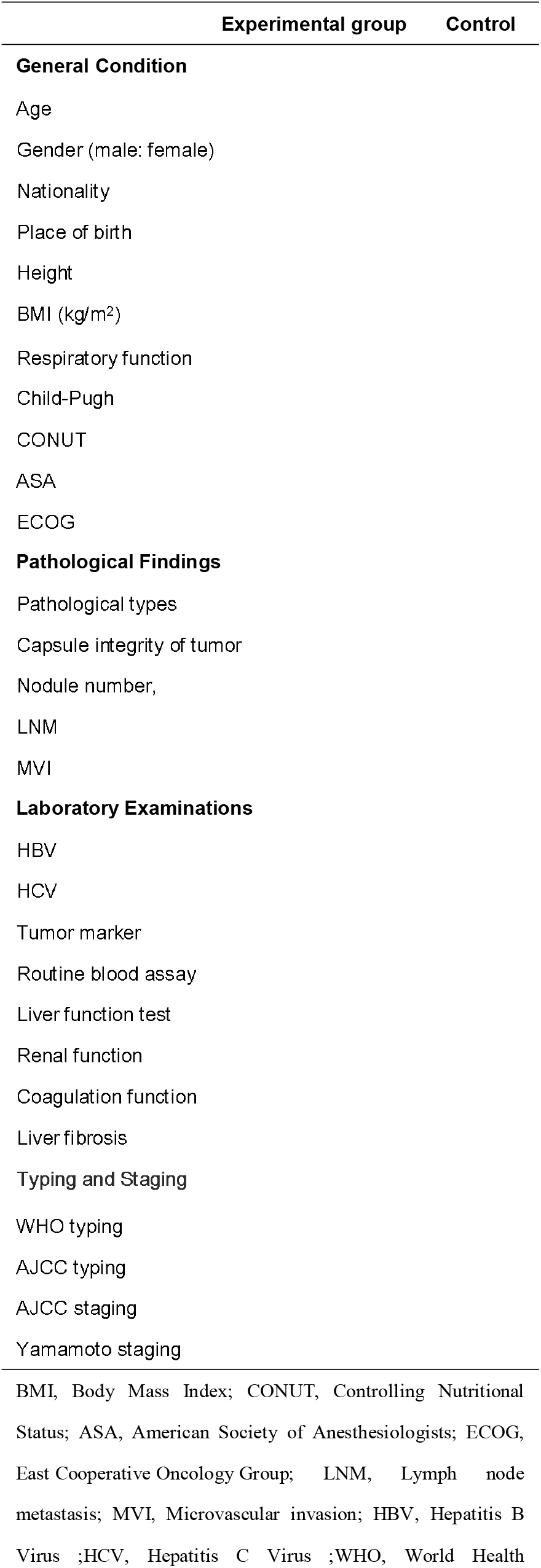
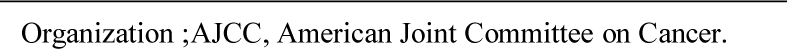
Demographic and clinical characteristics of the study participants

|  | Experimental group | Control |
| --- | --- | --- |
| **General Condition** |  |  |
| Age |  |  |
| Gender (male: female) |  |  |
| Nationality |  |  |
| Place of birth |  |  |
| Height |  |  |
| BMI (kg/m<sup>2</sup>) |  |  |
| Respiratory function |  |  |
| Child-Pugh |  |  |
| CONUT |  |  |
| ASA |  |  |
| ECOG |  |  |
| **Pathological Findings** |  |  |
| Pathological types |  |  |
| Capsule integrity of tumor |  |  |
| Nodule number, |  |  |
| LNM |  |  |
| MVI |  |  |
| **Laboratory Examinations** |  |  |
| HBV |  |  |
| HCV |  |  |
| Tumor marker |  |  |
| Routine blood assay |  |  |
| Liver function test |  |  |
| Renal function |  |  |
| Coagulation function |  |  |
| Liver fibrosis |  |  |
| Typing and Staging |  |  |
| WHO typing |  |  |
| AJCC typing |  |  |
| AJCC staging |  |  |
| Yamamoto staging |  |  |

BMI, Body Mass Index; CONUT, Controlling Nutritional Status; ASA, American Society of Anesthesiologists; ECOG, East Cooperative Oncology Group; LNM, Lymph node metastasis; MVI, Microvascular invasion; HBV, Hepatitis B Virus ;HCV, Hepatitis C Virus ;WHO, World Health

The inclusion criteria were: 1) Permanent residents over age 18 in the study area. 2) The preoperative diagnosis was resectable intrahepatic cholangiocarcinoma. 3) The patient has independent ability, can understand and voluntarily sign the informed consent form, and can complete the follow-up plan.

The exclusion criteria were:1) The postoperative diagnosis was not intrahepatic cholangiocarcinoma. 2) The patient failed to perform the operation as planned. 3) An unplanned disease or treatment that occurs to a patient. 4) Other situations that not suitable for continuing to participate in the trial. 5) The patient voluntarily withdrew from the trial.

### Informed consent

Patients participating in the experiments will all be given details of the study design and knowledge about intrahepatic cholangiocarcinoma. Patients were asked to sign an informed consent form at each stage of the study.

The trial’s informed consent is available as online supplemental material 1.

### Recruitment and randomisation

After fully understanding the experimental details and signing an informed consent form, the cholangiocarcinoma patients will be given a study number and enrolled into one of the study groups according to the randomization table: (1) Regional lymph node dissection group or (2) Extended lymph node dissection group. In the study, the research subjects were distinguished by the site code + number. Patients participating in the study were divided into two groups in a 1:1 ratio by the study coordinator according to a randomization table.

**Figure 1** displays the study design.

**Figure 1.**
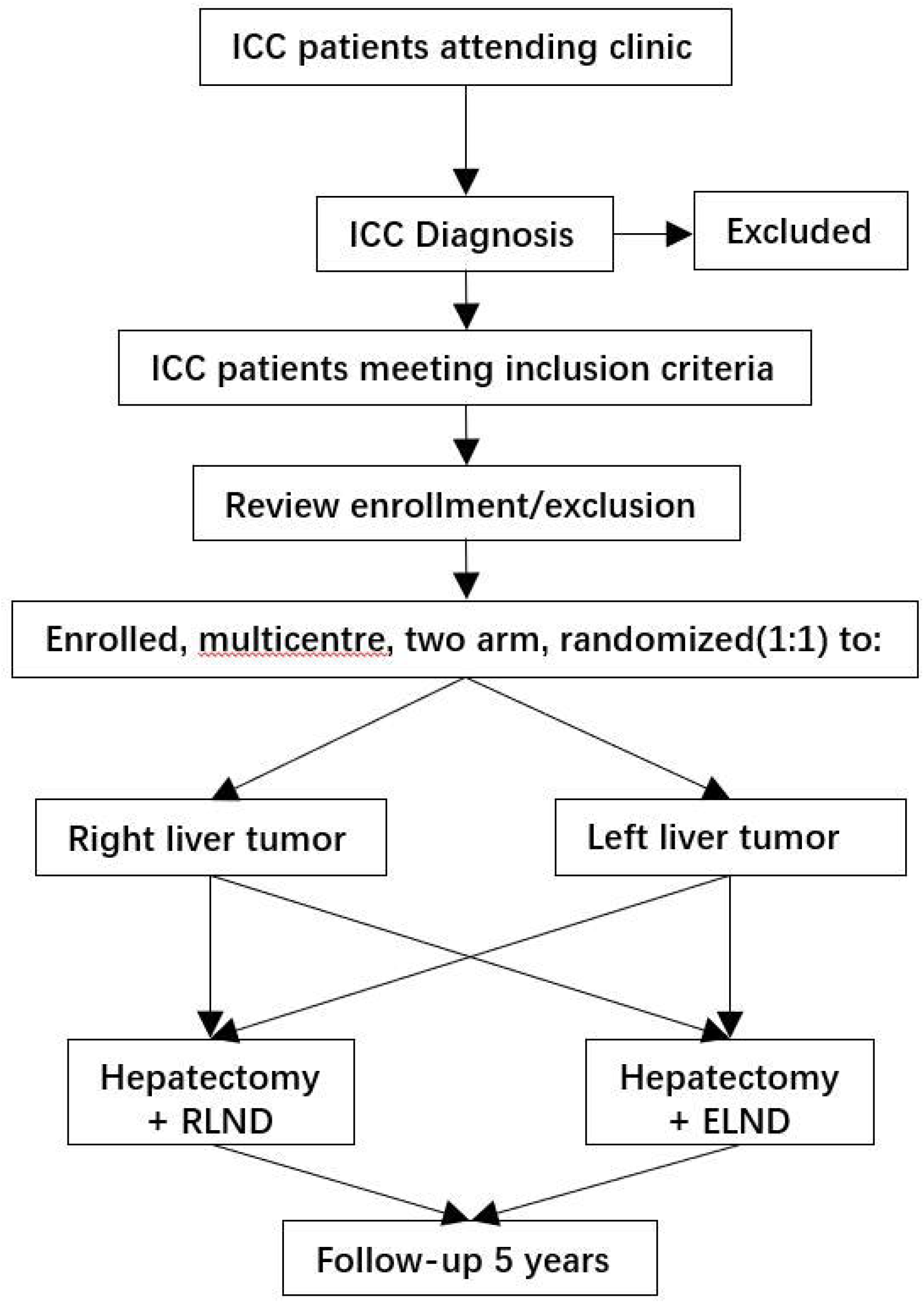
ICC LND trial design. ICC, Intrahepatic cholangiocarcinoma; RLND, Regional Lymph Node Dissection; Extended Lymph Node Dissection

### Blinding

All participants except patients, the study coordinator and the operative staff were kept blind throughout the study, and the operative staff did not participate in other aspects of the study. Unblinding is only envisaged in case of a medical emergency (in such case, the investigator on site will have to justify to the Data Safety Monitoring Board (DSMB) the need for unblinding).

### Interventions

#### regional lymph node dissection group

Hepatectomy + regional lymph node dissection (the 12th group lymph nodes). Surgical procedures: 1. Perform liver segment, hepatic lobe, hemi-hepatic or hepatic trilobe surgery according to the location of the primary tumor; 2. Skeletonization of the hepatoduodenal ligament; 3. The 12th group lymph nodes lymph nodes dissection.

#### extended lymph node dissection group

Hepatectomy + extended lymph node dissection (the 8th,12th and 13th group lymph nodes for the right lobe tumor; the 1st, 3rd,7th,8th and 12th lymph nodes for the left lobe tumor).

Surgical procedures: 1. Perform liver segment, hepatic lobe, hemi-hepatic or hepatic trilobe surgery according to the location of the primary tumor; 2. Skeletonization of the hepatoduodenal ligament; 3. The 8th,12th and 13th group lymph nodes dissection for the right lobe tumor; the 1st, 3rd,7th,8th and 12th lymph nodes for the left lobe tumor.

### Study outcomes

The primary outcome of the trial is disease-free survival (DFS). Secondary study outcomes include incidence of severe Clavien–Dindo complications (≥II), safety of operation, overall survival (OS), three-year survival rate, five-year survival rate, median mortality. tumor marker levels, liver function levels, blood routine levels, renal function levels.

### Sample size

Based on previous studies, we set the five-year survival rate after surgery for intrahepatic cholangiocarcinoma to be 15%, set bilateral α=0.05, β=0.2, power=1-β=0.8. The ratio of the number of cases in the two groups was 1:1. Calculated by PASS11, the required minimum sample size was 78 cases, with a total of 156 cases in the two groups.

The loses visit rate was set at 10%, and the minimum sample size of this study was 174 cases.

### Follow-up and measurements of outcomes

All information in the study will be recorded in the study specific Case Report Form (CRFs), the follow-up plan is shown in Table 3.

**Table 3.**
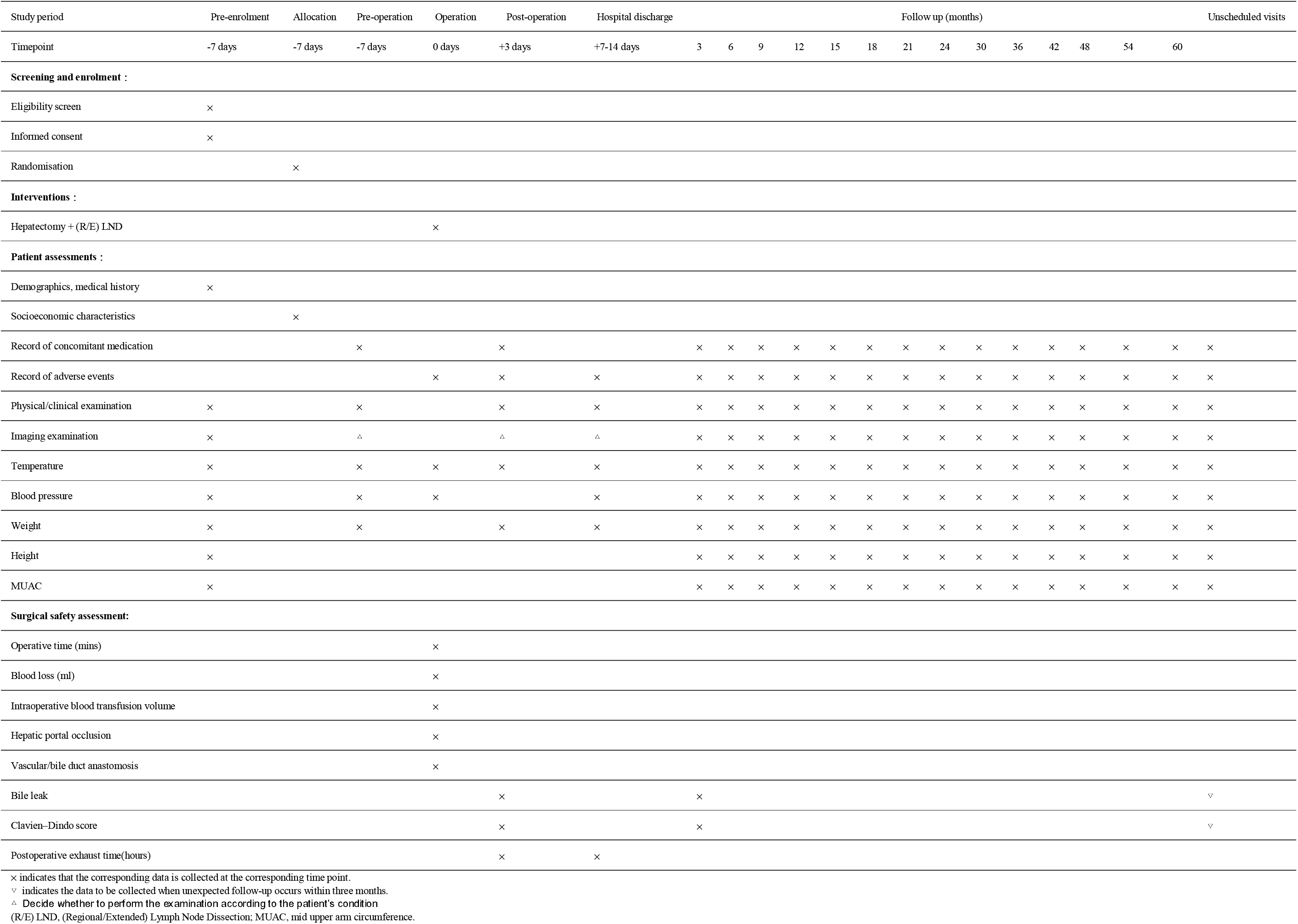
Schedule of enrolment, interventions and patient assessments

### Physical and clinical examination at enrolment

The physical examination of the ICC patients will include the following assessments: age, gender, nationality, place of birth, height, body mass index (BMI), medical history, respiratory function, Child-Pugh score, Controlling Nutritional Status(COUNT) score, American Society of Anesthesiologist(ASA) score, Eastern Cooperative Oncology Group(ECOG) score. Magnetic resonance imaging will be performed to confirm the diagnosis of intrahepatic cholangiocarcinoma and to evaluate the feasibility of surgical resection.

#### baseline biological samples

At enrolment, a blood sample will be collected for analysis of HAV, HBV, HCV, HEV, tumor marker, routine blood assay, liver function test, renal function, coagulation function, Cirrhosis.

#### follow-up and household visits

The patient will undergo a thorough examination and evaluation within 1 week of diagnosis. Participants will follow guidelines for intervention and are asked to report any discomfort to their physician. The patient will sign the informed consent before surgery and record the patient’s examination results. Blood routine examination, liver and kidney function and coagulation function were performed on the first postoperative day, and then every 3 days until discharge. Imaging examination was performed one week after operation to evaluate the recovery and complications. Patients are revisited every 3 months for the first 2 years after discharge and every 6 months thereafter until 5 years or death. Follow-up examination should include imaging examination, blood routine examination, tumor markers, and liver and kidney function, so as to assess the recurrence and progression of the tumor. If there is clinical suspicion of recurrence but no imaging evidence, further examination should be performed. Postoperative adjuvant therapy was performed according to the guidelines. Patients who cannot come to the hospital will be followed up by telephone and Electronic address.

### Laboratory tests

#### hepatitis virus detection

Peripheral blood HBsAg, HBsAb, HBeAg, HBeAb, HBcAb and HCVAb were detected with the corresponding ELISA kit according to the instructions on the first day of admission.

#### tumor marker detection

Tumor markers such as AFP, CA199 and CEA were detected by ELISA with corresponding kits before operation and at each follow-up.

#### routine blood detection

Peripheral blood was collected before operation, every 3 days after operation and during follow-up. After anticoagulation, routine blood test was performed by Hematology Analyzer.

#### blood biochemical examination

The liver and kidney function and coagulation function were measured before operation, every three days after operation and during follow-up.

#### liver fibrosis

Before operation, venous blood was collected and serum was isolated to detect the levels of hyaluronic acid(HA), laminin(LN), Procollagen peptide Type 3(PIIINP) and Collage Type IV(C-IV) in serum.

### Data management

Data collection was performed at each visit using a paper CRF form customized for the experiment, and the quality of study source documents and CRFs recorded data will be monitored by Clinical monitors following Good Clinical Practices. Data will be double-entered into the study database using OpenClinica open source software (V.3.14) for clinical data management at each study site. An automatic quality check is performed to ensure CRF integrity. The database system will automatically encrypt sensitive data to ensure data security and integrity, and include functions such as authorization, authentication and audit. Accompanying drugs registered in the database will be coded using the WHO Drug Reference List. This list uses the Therapeutic Chemical Classification System. History/current Medical conditions and adverse events will be coded using The Medical Dictionary for Regulatory Activities (MedDRA).

#### BOX 1

**Study outcomes**

**Primary endpoint**

The disease-free survival of patients after surgery (Time interval from the date of operation to tumor progression).

**Secondary endpoints**

Safety of operation.

Incidence of severe Clavien–Dindo complications (≥II).

Frequency and severity of adverse events.

Three-year survival rate.

Five-year survival rate.

Overall survival.

Median mortality.

### Analysis plan

The following analysis objectives have been identified: (A) Surgical effectiveness: This data compares disease-free survival, overall survival, 3-year survival, and 5-year survival in the two cohorts to compare the effectiveness of the two surgical methods for intrahepatic cholangiocarcinoma. (B) Surgical safety: The data were used to compare the operation time, intraoperative blood loss, intraoperative blood transfusion volume, whether to block the hepatic portal and the number and time of blocking, whether to perform vascular resection and anastomosis and the mode and time of anastomosis, the scope and number of lymph node dissection, the mode and time of biliary intestinal anastomosis, and the number of biliary intestinal anastomosis. incidence of serious postoperative complications, incidence of biliary fistula, and postoperative exhaust time between the two cohorts to compare the safety of the two surgical methods. (C) Tumor staging: This data compares the number of lymph nodes dissected and the positive rate of lymph nodes dissected by the two lymph node dissection procedures to determine whether expanded lymph node dissection contributes to accurate tumor staging.

### Patient and public involvement

Patients will not be directly involved in the research process, but each institution will have staff to maintain communication with patients to ensure that their reasonable demands are met. At the same time, research institutions should establish research committees and maintain communication to ensure the reliability, consistency and safety of the research.

### Ethics and dissemination

This study was approved by the the ethical review committee of the second affiliated hospital Zhejiang University school of medicine (2019-261). Trial results will be submitted to a peer-reviewed journal for publication within 1 year of study completion. The original project data will be published on the project website. No personal information about study participants will be released at any stage of the study.

## DISCUSSION

Intrahepatic cholangiocarcinoma is a malignant liver tumor with an incidence second only to hepatocellular carcinoma, and surgical resection is currently recognized as an effective treatment for intrahepatic cholangiocarcinoma.^[39]^

Studies have found that compared with hepatocellular carcinoma, intrahepatic cholangiocarcinoma is more prone to lymph node metastasis, and multiple studies have found that lymph node metastasis is an independent risk factor affecting the prognosis of intrahepatic cholangiocarcinoma. ^[40-43]^ Some studies have also pointed out that lymph node metastasis of intrahepatic cholangiocarcinoma is an intermediate step in distant metastasis. A study of intrahepatic cholangiocarcinoma reported that intrahepatic cholangiocarcinoma may have skip metastasis, half of the patients had direct second-stop lymph node metastasis. ^[44]^ Based on the above research inferences, thorough lymph node dissection is helpful for accurate staging of intrahepatic cholangiocarcinoma and is beneficial to prevent distant metastasis of the seed tumor. These conjectures were also confirmed in subsequent studies. ^[2]^ Therefore, lymph node dissection during radical resection of intrahepatic cholangiocarcinoma has become the consensus of major guidelines.

However, the scope of lymph node dissection is still controversial in different guidelines and studies. ^[15]^ Some studies have suggested that extended lymph node dissection can prolong the operation time, increase the intraoperative risk and may increase the incidence of serious complications such as biliary fistula, and has no significant effect on improving the prognosis of patients. ^[45,46]^ Other studies have found that expanded lymph node dissection can help to completely remove the tumor, prevent tumor recurrence, accurately judge tumor staging, and guide postoperative adjuvant therapy. It is beneficial to prolong the patient’s survival time and improve the quality of life. ^[24,27,47-49]^

At present, the studies on the comparison between regional lymph node dissection and expanded lymph node dissection in patients with intrahepatic cholangiocarcinoma are mostly retrospective small-sample studies with a long history (> 10 years). In recent years, the innovation of laparoscopic technology, robotic technology and other surgical techniques have shortened the operation time, improved the quality of surgery and reduced the surgical injury. Therefore, under the current background, it is necessary to carry out a multicenter, prospective, large-sample randomized controlled study to compare the efficacy of regional lymph node dissection and expanded lymph node dissection for intrahepatic cholangiocarcinoma.

In conclusion, in order to provide a reliable basis for standardizing the extent of lymph node dissection during radical resection of intrahepatic cholangiocarcinoma, we will implement a multicenter prospective randomized controlled study. To compare the efficacy of regional lymph node dissection and extended lymph node dissection in patients undergoing radical resection for intrahepatic cholangiocarcinoma, and to compare the safety and prognostic effects of the two procedures.

## Data Availability

This is an open access article distributed in accordance with the Creative Commons Attribution NonCommercial (CC BY- NC 4.0) license, which permits others to distribute, remix, adapt, build upon this work non- commercially, and license their derivative works on different terms, provided the original work is properly cited, appropriate credit is given, any changes made indicated, and the use is non-commercial. See: http://creativecommons.org/licenses/by-nc/4.0/

## Contributors

LQ and FY are joint first authors. Conceived and designed the study: LQ and LJT. Gave inputs to protocol methodology: LQ, FY, XL, JY, YY, GY, ZX and LJ. Wrote the first draft of the manuscript: LQ and FY. Wrote sections on sample size, data management and analysis plan: LQ, XL and JY. Wrote, reviewed and approved the manuscript: all authors.

## Disclaimer

The project funders do not have any role on study design, data collection and analysis.

## Ethical approval

This study was approved by the ethical review committee of the second affiliated hospital Zhejiang University school of medicine (2019-261)

## Patient consent for publication

Not applicable.

## Provenance and peer review

Not commissioned; externally peer reviewed.

